# Has the implementation of time-based-targets for emergency department length-of-stay influenced the quality of care for patients? A systematic review of qualitative literature

**DOI:** 10.1101/2021.01.03.21249171

**Authors:** Katie Walker, Bridget Honan, Daniel Haustead, David Mountain, Vinay Gangathimmaiah, Ella Martini, Roberto Forero, Rob Mitchell, Greg Tesch, Ian Bissett, Peter Jones, Yusuf Nagree, Paul Middleton, Danny Liew

## Abstract

**Background:** Time-based-targets for emergency department length-of-stay were introduced in England in 2000; followed by Canada, Ireland, New Zealand, and Australia after emergency department crowding was associated with poor quality of care and increased mortality.

**Objectives:** The aim of the systematic review was to evaluate qualitative literature to investigate how implementing time-based-targets for emergency department length-of-stay has influenced the quality of care of patients.

**Methods:** Systematic review of qualitative studies that described knowledge, attitudes to or experiences regarding a time-based-target for emergency department length-of-stay. Searches were conducted in Cochrane library, Medline, Embase, CInAHL, Emerald, ABI/Inform, and Informit. Individual studies were evaluated using the Critical Appraisal Skills Programme tool. Individual study findings underwent thematic analysis. Confidence in findings was assessed using the Confidence in the Evidence from Reviews of Qualitative research approach.

**Results:** The review included thirteen studies from four countries, incorporating 617 interviews. Themes identified were: quality of care, access block and overcrowding, patient experience, staff morale and workload, intrahospital and interdepartmental relationships, clinical education and training, gaming, and enablers and barriers to achieving targets. The confidence in findings is moderate or high for most themes. More patient and junior doctor perspectives are needed.

**Conclusions:** Emergency time-based-targets have impacted on the quality of emergency patient care. The impact can be both positive and negative and successful implementation depends on whole hospital resourcing and engagement with targets.

**Funding:** The Australasian College for Emergency Medicine provided administrative support for the study, no funding was received.

**Registration:** PROSPERO CRD42019107755 (prospective)

## INTRODUCTION

Long waits for assessment and admission to hospital from emergency departments are a problem around the world. Long waits contribute to ED crowding and adverse outcomes for patients, including excess mortality ^1-3^. To address these issues, various jurisdictions have introduced time-based-targets for ED length-of-stay. Time-based-targets were first implemented in the United Kingdom in 2004, following the 2000 National Health System plan that stated:

> “*By 2004 no-one should be waiting more than four hours in accident and emergency from arrival to admission, transfer or discharge. Average waiting times*…*will fall as a result to 75 minutes. By then we will have ended inappropriate trolley waits for assessment and admission*.*”*^*4*^

Following this lead, governments introduced time-based-targets for emergency stays in Australia (4 and 8 hours)^5^ New Zealand (6 hours)^6^, Canada (8 hours)^7^, and Ireland (6 hours). A description of the targets in different jurisdictions including incentives is shown in Table 1.

**Table 1.** Time-based targets for emergency length-of-stay in different jurisdictions

| Jurisdiction | Target time / Name | Target thresholds and dates | Target incentives | Source data |
| --- | --- | --- | --- | --- |
| England | Four hours<br>Four Hour Rule (FHR) | Target pledged in July 2000: "By 2004 no-one should be waiting more than four hours in accident and emergency from arrival to admission, transfer or discharge."<sup>4</sup><br>The initial target announced was 100% but reduced to 98% at the behest of clinicians in 2004 prior to full implementation.<br>In 2010, the target was relaxed to 95% of all patients within four hours.<br>The current target, as of October 2019, is for at least 95% of all patients should be admitted, transferred or discharged within four hours from arrival in the ED. | In each of the five fiscal quarters from January 2005 hospitals would receive a lump-sum grant of £100,000 if the percent of patients treated within 4 hours across the entire quarter rose above a threshold (total of £500,000 for each ED if sequential targets met).<br>NHS Trust performance publicly reported and contributed to Trust 'Star Rating' from 2003.<br>Strong ministerial pressure to meet the target | <sup>4 8 9</sup> |
| Ontario, Canada | Four hours (low acuity target)<br>Eight hours (high acuity target)<br>Wait Time Targets | In April 2008, the Ontario Ministry of Health launched the ED Wait Time Strategy<br>High acuity target: no more than 8 hours in ED (from time of registration or triage to completion of visit) for 90% of patients<br>Low acuity target: no more than 4 hours in ED (from time of registration or triage to completion of visit) for 90% of patients | Pay-for-performance program provided annual financial incentives to hospitals for improved performance on ED LOS targets<br>Hospital performance publicly reported via Health Quality Ontario website | <sup>7 10 11</sup> |

| **Jurisdiction** | **Target time / Name** | **Target thresholds and dates** | **Target incentives** | **Source data** |
| --- | --- | --- | --- | --- |
| New Zealand (NZ) | Six hours<br>Shorter Stays in Emergency Departments Target (SSED) | In 2008, the NZ Ministry of Health announced a national health target of 95% of patients to be admitted, discharged or transferred from an ED within 6 hours. This target was implemented from July 2009. | Ministerial pressure on DHB management, league tables published in newspapers until 2017. No financial incentives | <sup>12-13</sup> |
| Ireland | Six hours | Graduated reduction from a 24-hour target in 2006 for admission from ED. A 12-hour target was set in October 2007 aiming for six hours in January 2009. Threshold not specified but implied 100% at this time (later shifted to 95% in June 2012), applied to all ED patients | Not clear. Extra money was provided by the government to improve emergency care in a 10-point plan but this did not appear to be linked to target performance | <sup>14-17</sup> |
| Australia:<br>Western<br>Australia<br>(WA) | Four Hours<br>Initially Four-Hour Rule Program (FHRP)<br>Later National Emergency Access Target (NEAT) | FHRP implemented in stages, commencing with four tertiary hospitals in April 2009, followed by three secondary hospitals in October 2009 and remaining regional hospital in stage three.<br>The FHRP had a staggered increase target percentage of patients arriving at EDs seen and admitted, discharged or transferred within four hours) starting at 85% in April 2010 and increasing to 98% in April 2011. After review, the April 2011 target for three of the four major hospitals was revised to 85%. One paediatric hospital retained its 98% target.<br>In April 2011 NEAT was applied at a federal level with objective to apply target to 90% of all patients by 31 December 2015.<br>In January 2016, West Australian Emergency Access Target was established and required that 90% of all patients presenting to a public hospital ED will be seen and admitted, transferred or discharged within four hours. | WA received a 'one-off payment of \$75.285 million to implement specific initiatives to manage emergency demand' under a Commonwealth Government National Partnership Agreement project payment, of which \$56 million was allocated to the FHRP.<br>Between 2012- 2015, WA was eligible for Annual Reward Funding from the Commonwealth government upon meeting NEAT, to a value of \$5.3 million annually. Reward funding for meeting NEAT ceased in July 2015<br>MyHospitals website started publicly reporting performance in 2011-2012 | 18-20 |
| Australia: Victoria | Four Hours<br>National Emergency Access Target (NEAT) | In April 2011 NEAT was applied at a federal level with objective to apply target to 90% of all patients by 31 December 2015.<br>Staggered increase over four year period, aiming to apply to 70% of patients in 2012, 75% of patients in 2013, 81% of patients in 2014 and 90% of patients in 2015.<br>After cessation of NEAT in 2015, the Victoria DHHS set a new target of 81% of patients who present at public hospital EDs to be admitted to the hospital for treatment, referred to another hospital for treatment, or discharged within four hours. | Prior to NEAT, the Victorian Department of Human Services (DHS) determined a benchmark that 80% of non-admitted emergency patients patients should have a LOS <4 hours, and 80% of emergency patients who were admitted to an inpatient bed should be transferred within 8 hours<br>Between 2012- 2015, Victoria was eligible for Annual Reward Funding from the Commonwealth government upon meeting NEAT, to a value of \$12.4 million annually. Reward funding for meeting NEAT ceased in July 2015<br>MyHospitals website started publicly reporting performance in 2011-2012 | 19- 22 |
| Australia: Queensland | Four Hours<br>National Emergency Access Target (NEAT) | In April 2011 NEAT was applied at a federal level with objective to apply target to 90% of all patients by 31 December 2015.<br>Staggered increase over four year period, aiming to apply to 70% of patients in 2012, 77% of patients in 2013, 83% of patients in 2014 and 90% of patients in 2015.<br>Following cessation of NEAT, Queensland froze its target at 83% while undertaking a review. Based on the review, the Queensland Emergency Access Target (QEAT) applying to 80% of all patients presenting to ED came into effect in 2016 or 2017. | Between 2012- 2015, Queensland was eligible for Annual Reward Funding from the Commonwealth government upon meeting NEAT, to a value of \$10.4 million annually. Reward funding for meeting NEAT ceased in July 2015<br>MyHospitals website started publicly reporting performance in 2011-2012 | 23 24 |
| Australia:<br>New South Wales (NSW) | Four Hours<br>National<br>Emergency<br>Access Target<br>(NEAT) | In April 2011 NEAT was applied at a federal level with objective to apply target to 90% of all patients by 31 December 2015.<br>Staggered increase over four year period, aiming to apply to 69% of patients in 2012, 76% of patients in 2013, 83% of patients in 2014 and 90% of patients in 2015.<br>Following cessation of NEAT, reverted to a target of 81%, referred to as Emergency Treatment Performance (ETP). | Between 2012- 2015, NSW was eligible for Annual Reward Funding from the Commonwealth government upon meeting NEAT, to a value of \$15.9 million annually. Reward funding for meeting NEAT ceased in July 2015<br>MyHospitals website started publicly reporting performance in 2011-2012 | 19 20 25-28 |
| Australia:<br>Tasmania | Four Hours<br>National<br>Emergency<br>Access Target<br>(NEAT) | In April 2011 NEAT was applied at a federal level with objective to apply target to 90% of all patients by 31 December 2015.<br>Staggered increase over four year period, aiming to apply to 72% of patients in 2012, 78% of patients in 2013, 84% of patients in 2014 and 90% of patients in 2015.<br>After cessation of NEAT, Tasmania implemented a target of 80% of ED LOS for all emergency patients, in 2016. In 2018, the target was increased to “not less than 90%” to be achieved by 2022. | Between 2012- 2015, Tasmania was eligible for Annual Reward Funding from the Commonwealth government upon meeting NEAT, to a value of \$1.1 million annually. Reward funding for meeting NEAT ceased in July 2015<br>MyHospitals website started publicly reporting performance in 2011-2012 | 19 20 29 30 |
| Australia:<br>South<br>Australia (SA) | Four Hours<br>National<br>Emergency<br>Access Target<br>(NEAT) | In April 2011 NEAT was applied at a federal level with objective to apply target to 90% of all patients by 31 December 2015.<br>Staggered increase over four year period, aiming to apply to 67% of patients in 2012, 75% of patients in 2013, 82% of patients in 2014 and 90% of patients in 2015.<br>After cessation of NEAT, there does not appear to have been a specifically defined target for ED LOS in SA, however the SA Health Department does publish an ED dashboard which reported the proportion of patients with an ED LOS < 4 hours. | Between 2012- 2015, SA was eligible for Annual Reward Funding from the Commonwealth government upon meeting NEAT, to a value of \$3.5 million annually. Reward funding for meeting NEAT ceased in July 2015<br>MyHospitals website started publicly reporting performance in 2011-2012 | 19 20 31 |
| Australia:<br>Australian<br>Capital<br>Territory<br>(ACT) | Four Hours<br>National<br>Emergency<br>Access Target<br>(NEAT) | In April 2011 NEAT was applied at a federal level with objective to apply target to 90% of all patients by 31 December 2015.<br>Staggered increase over four year period, aiming to apply to 64% of patients in 2012, 73% of patients in 2013, 81% of patients in 2014 and 90% of patients in 2015.<br>After cessation of NEAT, ACT Department of Health has continued to set targets for ED LOS < 4 hours in annual reports, from 69% in 2015-2016, to 90% in 2017-2018. | Between 2012- 2015, ACT was eligible for Annual Reward Funding from the Commonwealth government upon meeting NEAT, to a value of \$0.8 million annually. Reward funding for meeting NEAT ceased in July 2015<br>MyHospitals website started publicly reporting performance in 2011-2012 | 19 20 32 |
| Australia:<br>Northern<br>Territory (NT) | Four Hours<br>National<br>Emergency<br>Access Target<br>(NEAT) | In April 2011 NEAT was applied at a federal level with objective to apply target to 90% of all patients by 31 December 2015.<br>Staggered increase over four years, aiming to apply to 69% of patients in 2012, 78% of patients in 2013, 84% of patients in 2014 and 90% of patients in 2015.<br>After cessation of NEAT, the NT government announced a target of 78% of ED presentations to have a LOS < 4 hours. | Between 2012- 2015, the NT was eligible for Annual Reward Funding from the Commonwealth government upon meeting NEAT, to a value of \$0.6 million annually. Reward funding for meeting NEAT ceased in July 2015<br>MyHospitals website started publicly reporting performance in 2011-2012 | 20 33 |

Concerns have been raised about the potential for unintended effects of targets and gaming *‘hitting the target but missing the point’*^34-36^. As a result, several jurisdictions have relaxed or de-emphasised these targets^37 38^.

Since the last quantitative systematic review in 2010^39^, many quantitative and qualitative studies have been published reporting outcomes for patients from different countries with various time-based-targets for emergency length-of-stay. To date, there hasn’t been any synthesis of qualitative research on the impact of time-based-targets, hence the impact of targets across settings remains unknown. Compared to individual qualitative studies, a systematic review improves external generalisability of the qualitative research to other settings. This should better inform the complex and often political discussion about whether time-based target policy instruments should be implemented or maintained.

The aim of the study is to evaluate qualitative evidence to investigate how implementing time-based-targets for emergency medicine patient length-of-stay has influenced the quality of care of patients.

## METHODS

A research team, comprising emergency physicians, consumers, academic researchers, and non-emergency specialists was convened at the request of ACEM in 2018/19. The research question was: How has the implementation of time-based-targets for emergency length-of-stay impacted on the quality of patient care? A secondary question that arose during the study was: What are the barriers and enablers to implementing time-based-targets in emergency medicine?

Ethics approval was not required. The study was registered prospectively at PROSPERO (CRD42019107755). The study protocol is available at https://www.crd.york.ac.uk/prospero/display_record.php?RecordID=107755.

### inclusion criteria

Original qualitative or mixed methods research studies were eligible for inclusion if they described a state, regional or national time-based-target for emergency length-of-stay; were published between 2000 and 2019 and reported knowledge, attitudes or experiences regarding the target. Included papers, in any language, needed to describe emergency department patients and the perspectives of stakeholders who interacted with emergency medicine before and after the introduction of the targets. Data could be acquired through interviews or focus groups. Review articles, surveys, simulation studies, secondary reports of other studies, news articles, letters and editorials were excluded.

### search strategy

Structured searches were undertaken in the Cochrane library, Medline, Embase, Cumulative Index to Nursing, and Allied Health Literature, Emerald, Abstracted Business Information/Inform and Informit (final search on October 23rd 2019). A search strategy was developed using text words and index terms related to synonyms for time-based targets, emergency departments, and any term related to outcomes, quality or evaluation. Search strategies are in Appendix 1. Authors were consulted regarding unpublished work and hand searches were performed.

### study screening

Abstracts were placed in a bibliographic management tool (Mendeley)^40^ and duplicates were removed. Remaining abstracts were uploaded to Covidence systematic review software^41^. Full texts screening followed abstract screening for all abstracts potentially eligible for inclusion. Two reviewers screened each abstract and full text article, (blinded to the second reviewer assessment). Inter-rater agreement during initial full-text screening was measured using a kappa statistic. Disagreements were resolved either by a consensus discussion between reviewers or a third reviewer. Reasons for excluding publications were recorded during full-text screening.

### data extraction

The following data were extracted: title; journal; date of publication; sponsorship source; country; setting; lead author name, institution, and contact details; methodology (design, when data was collected, analysis and synthesis); participant inclusion and exclusion criteria; recruitment method; source of data, baseline characteristics of participants and sites; type of target; details of the target; date of target introduction; incentives to meet targets and outcomes from the study. Illustrative quotes from respondents were extracted from each included study. Each screening reviewer verified the extracted data.

### quality appraisal of individual studies

Individual study demographic and methods data were collected using a data extraction form in Covidence. Data regarding study outcomes, quality and risk of bias were collected on a data extraction form, based on the Critical Appraisal Skills Program (CASP)^42^ requirements for qualitative study evaluation. Domains assessed were: aims, methodology and design, recruitment, data collection, analysis, outcomes, and an assessment of overall risk of bias. The template is available in Appendix 2. Two reviewers independently extracted data and one reviewer combined the results. Disagreements were resolved through discussion or by a third reviewer. No studies were excluded from the review based on this assessment.

### data synthesis

Two emergency physician reviewers individually identified all the key outcomes for each study, and interpreted each outcome as positive, negative or neutral, with regards to the impact of the targets on quality of care. Reviewers weren’t provided with definitions of positive/negative/neutral. A thematic synthesis approach was used to analyse and synthesise individual study outcomes. Reviewers iteratively classified all the findings from the studies into themes. Regular meetings were held throughout the data synthesis process for reviewers to compare and agree on descriptive/emerging themes and interpretations. Quotations were selected to illustrate each theme.

### assessing confidence in the review findings

Assessment of confidence in the finding was undertaken using the Confidence in Evidence from Reviews of Qualitative Research (CERQual)^43^, which applies the Grading of Recommendations, Assessment, Development and Evaluations (GRADE) approach to qualitative studies^44 45^. CERQual assesses each review finding against four domains: methodological limitations^46^, relevance^47^, coherence^48^ and adequacy of data^49^. Reviewers categorised confidence in each finding as high, moderate, low or very low, based on the assessments for each CERQual component^50^, with equal weight given to each component. High confidence in a synthesised finding indicates that it is a reasonable representation of time-based targets. If there are concerns with regard to any of the above four domains, then confidence in the finding is downgraded. Where the domains overlap, reviewers avoided downgrading a review finding twice for the same concern across domains. Two reviewers individually undertook the assessment of confidence in findings using the CERQual approach.

## RESULTS

### study selection

The flow of studies through the review is described in Figure 1. Initial inter-observer full text inclusion agreement was 98/121 (81%), κ=0.60 (95% CI 0.45-0.74), indicating moderate agreement.

**Fig 1.**
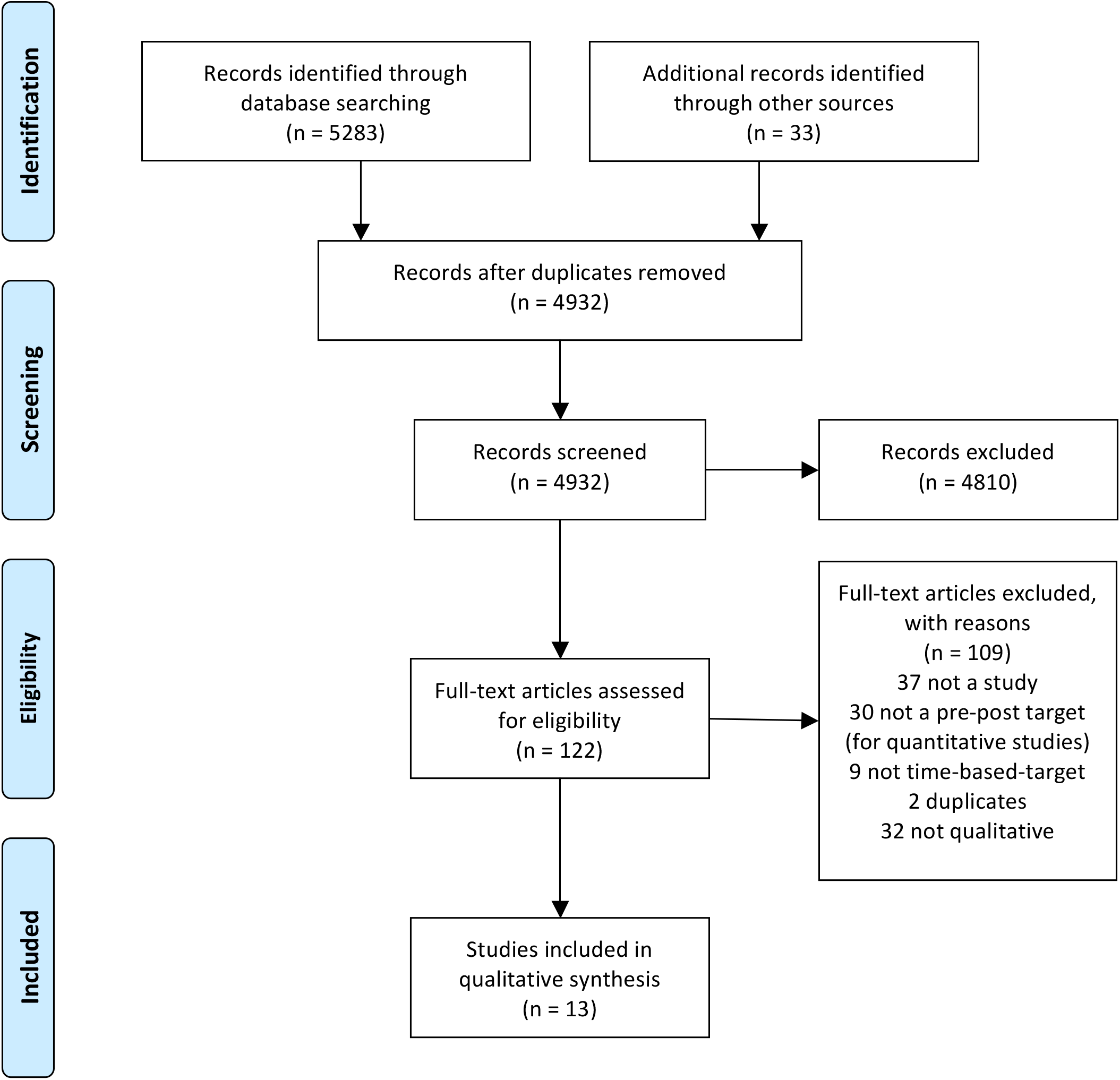
PRISMA Flow Diagram, ED time-based-targets, flow through qualitative Systematic review. *From:* Moher D, Liberati A, Tetzlaff J, Altman DG, The PRISMA Group (2009). *P*referred *R*eporting *I*tems for *S*ystematic Reviews and *M*eta-*A*nalyses: The PRISMA Statement. PLoS Med 6(7): e1000097. doi:10.1371/journal.pmed1000097 **For more information, visit www.prisma-statement.org**.

### study characteristics

Thirteen studies were included in the review, and their characteristics including the setting, perspectives of respondents and the target intervention are presented in Table 2.

**Table 2.** Summary of Settings, Perspectives and Interventions for included studies

| Paper | Setting | Perspectives | Intervention | Outcomes |
| --- | --- | --- | --- | --- |
| Forero, 2019<sup>51</sup> | 16 metropolitan hospitals in four Australian states and territories: Western Australia (WA), Queensland (QLD), New South Wales (NSW), and the Australian Capital Territory (ACT) | 119 interviews conducted among ED staff between 2015 and 2016. Study participants included department directors, physicians, nurses and administrative employees working in the ED. No senior management. | WA piloted the first time-based target in Australia, called the Four-Hour Rule Program from 2009. All Australian states/territories had some form of 4-hour target from April 2011 onwards. See Table 1 for more details. | 1. Positive social factors: whole of hospital approaches improved patient flow, executive buy-in improved in some hospitals, some staff-patient communication increased 2. Negative social factors: stress and bullying increased, morale, inter-disciplinary relationships and teamwork deteriorated, some staff-patient communication deteriorated 3. ED operations: Staff shortages increased with deteriorating working environments; one-call and direct admission policies increased; IT systems changed; size and capacity of EDs increased 4. Education and training deteriorated for most 5. Changes weren't sustained and after an initial plateau, most failed |
| Forero, 2019<sup>52</sup> | As above | As above | As above | 1. Positive personal experiences of stress and morale: more efficient decision-making, less access block and improved intra-ED teamwork improved stress levels and morale 2. Negative personal experiences of stress and morale: work-load and administrative burden increased, junior doctors had to make decisions faster, change was chaotic and confusing for some; bullying increased; ambulances handed over patients before EDs had capacity; adversarial relationships developed between doctors and nurses; working conditions were strained and sick leave increased 3. Intergroup dynamics: intra-ED communication improved; inter-disciplinary relationships deteriorated 4. Hospital executive support sometimes improved; whole hospital approaches improved ED patient flow 5. Some patients experienced better communication, some worse |
| Nahidi, 2019<sup>53</sup> | As above | As above | As above | 1. Enablers and barriers: Factors that enabled successful implementation of TBT included whole-of-hospital and top-down management buy-in, changes to staffing, processes and models of care, and financial investment. Barriers to achieving TBT included lack of inpatient capacity, lack of ED authority regarding admissions, insufficient support from logistics or other areas of the hospital, increasing ED attendances (also increasingly complex patients) and lack of motivation of ED staff regarding working towards the target. 2. Overcrowding: TBT could either improve or worsen ED overcrowding, and that TBT could be associated with improvements or worsening of quality of care. 3. Gaming occurred. 4. Clinical education and training suffered. |
| Nahidi, 2019<sup>54</sup> | As above | As above | As above | 1. Time-based-targets required management changes, education and staff training as well as financial incentives<br>2. Pursuing TBT led to changes in staffing, models of care and processes, inter-departmental relationships, infrastructure and IT<br>3. Barriers to achieving TBT included lack of inpatient capacity, lack of ED authority regarding admissions, insufficient support from logistics or other areas of the hospital, increasing ED attendances (also increasingly complex patients) and a lack of motivation of ED staff to work towards the target. |
| Stokes, 2011<sup>55</sup> | Fremantle Hospital, Princess Margaret Hospital, Royal Perth Hospital and Sir Charles Gairdner Hospital, which were the first hospitals to implement targets in Western Australia and Australia. | 316 interviews (70 senior nurses, 59 consultant physicians, 51 junior doctors, 38 patient support staff, 27 allied health, 34 clinical leads, 13 4hr-rule implementation staff, 11 business staff, 9 directors, 6 patient liaison nurses, 4 staff nurses, 3 Australian Medical Association representatives, Australian college of surgeon representatives, members of public, health consumer agencies (7% participants)). Unclear when the interviews took place; the report was published in December 2011. | Prior to the introduction of NEAT, Western Australia piloted targets. This was implemented in stages, commencing with 4 tertiary hospitals in April 2009, followed by 3 secondary hospitals in October 2009. See Table 1 for more details. | 1. Poor engagement with medical staff. 2. Solutions to poor TBT performance related to whole-of-hospital changes, new processes and models of care. 3. Increased time pressure on staff. 3. Reduced training opportunities for junior doctors. 4. Improved patient satisfaction. 5. Risks to safety and quality of care. 6. Gaming and inappropriate admissions occur in response to TBT |
| Leggat, 2016<sup>56</sup> | Single metropolitan hospital in Melbourne, Australia | Managers and clinicians in the ED and hospital (9 managers, 9 nurses, 7 doctors and 1 allied health professional. Interviews being conducted in 2012 and 2013 (4 years after the intervention commenced). | Prior to NEAT, the Victorian Department of Human Services (DHS) determined a benchmark that 80% of non-admitted emergency patients should stay less than 4 hours, and 80% of emergency patients who were admitted to an inpatient bed should be transferred within 8 hours. See Table 1 for more details. | 1. Coordination, collaboration and team-work needed was hard to achieve. 2. Staff found “production type” processing of patients compromised their care of individual patients. |
| Tenbenschel, 2020<sup>57</sup> | 4 case study hospitals in New Zealand (NZ). Two large urban centres with district population >400,000; 1 medium-sized urban centre serving regional population with district population | 68 interviews (36 clinicians - allied health, nursing, medical; 10 clinical managers; 22 non-clinical managers; 29 ED staff; 39 non-ED staff). Interviews were conducted in two rounds in 2011 and 2012, which was 18 months and 3 years | The Shorter Stays in Emergency Department policy was launched by the NZ government in 2009 and had a target of target length-of-stay of 6 hours for 95% of all ED patients. See Table 1 for more details. | 1. Decanting to stop timing patient stays: allows patients to stay in the ED until the morning; delays decisions regarding appropriate inpatient units, hence delaying access to care and diverting attention to other patients 2. Motivations for gaming vary from one site to another 3. Gaming knowledge and abilities increase over time |
|  | 200-400,000 and one small urban centre with a population of 100-200,000. | respectively, after implementation of the targets. |  |  |
| Tenbensen, 2017<sup>58</sup> | As above | As above | As above | 1. In order to achieve TBT, hospitals required increased staffing, infrastructure including physical bed capacity, IT and communication systems. 2. Strong commitment from hospital executive and “whole of hospital” response was required to achieve TBT. 3. Gaming occurred to meet the TBT. 4. TBT was associated with antagonism between ED and other departments. 5.TBT was associated with improved patient flow |
| Tenbensen, 2016<sup>59</sup> | As above | As above | As above | 1. Increased pressure on ED and ward staff and reduced morale. 2. Exacerbation of tensions between ED and inpatient departments. 3. Positive TBT experience was associated with whole-of-hospital approach and improved systems/processes. 4. Clear evidence of gaming |
| Weber, 2011<sup>60</sup> | 9 Trusts in England | Participants were the Lead Clinician, Head Nurse and Business manager of each ED. Interviews were conducted between June - August 2008. | England’s National Health Service mandated that 98% of ED patients should stay less than 4 hours in ED from January 2005. See Table 1 for more details. | 1. Need for whole-of-hospital engagement and responsibility for emergency care. 2. ED nurses experienced increased autonomy but also increased workload and stress. 3. Improved patient flow and reduced trolley waits. 4. Critical care patients spent less time in ED, which could be perceived as positive or negative. 5. Reduced time for training junior doctors |
| Mortimore<sup>61</sup> | Single District General Hospital in England | Participants were 3 charge nurses, 3 staff nurses and 3 health care assistants, and were required to have worked in an ED for >5 years to ensure they had experience of waiting times prior to the implementation of the 4-hour target. Unclear when the interviews took place. | See above | 1. TBT associated with improvement in patient flow and journey; increased patient satisfaction. 2. Increased staff workload and reduced morale. 3. Distorted clinical priorities. 4. Unintended consequences: reduced training opportunities, increased attendances due to decreased queues, increased inpatient bed occupancy. 5.Need for increased axillary services for ED (diagnostics, pharmacy) |
| Vezyridis, 2014<sup>62</sup> | UK Hospital with 146,000 ED attendances annually. | The 28 participants in this study (23 female and 5 male) included the system administrator, the change manager, 2 Emergency Department Assistants, the operational services coordinator, 4 Emergency Nurse Practitioners, 4 charge nurses and 15 staff nurses. Interviews were conducted over 8 months (April - November 2008). | See above | 1. Physical space and layouts changed for the better (privacy, work-flow, security) 2. Some patients couldn't be categorised and placed in an appropriate area. 3. Time targets became more important than patient care 4. Information technology improved whole department awareness of patients and their times in the ED 5. New roles allowed medical task shifting 6. Some communications became combative in efforts to improve flow, perceived as bullying and harassment 7. Some patients were diverted to alternate services (perceived positively and negatively) |
| Rotteau, 2015<sup>63</sup> | 10 hospitals in Ontario, ranging in average annual ED volume from 27,967 - 71,523 presentations; proportion of admitted patients ranging from 5.1% - 19.8%. | 51 respondents from 10 different hospitals (out of 36), all from the "implementation team" but in a range of different roles (executive sponsor, physician lead, team lead, mix of ED and inpatient teams). Interviews were conducted between November 2010 and September 2011. | Ontario Emergency Department Wait Time Strategy implemented in 2008 with target times of 8 hours for high acuity patients and 4 hours for low acuity patients. The Ontario ED Process Improvement Program was launched in three waves between March 2009 and | 1. Local context: change was easier with visible leadership support; lack of substantial process improvement experience, lack of means to repeat measurements hindered implementation 2. Relationships between the improvement team and clinical teams was important; multi-professional representation was important; coaches could be a positive or negative influence 3. Staff engagement: enhanced when changes were patient centred; effective leadership was needed to engage sceptical physicians; frontline staff could be resistant to change 4. Success and sustainability: some facilities accepted the need for permanent and long-term changes; sustainability is difficult and usually behaviour returns to baseline |
|  |  |  | December 2010<br>within 36 hospitals<br>across Ontario. See<br>Table 1 for more<br>details. |  |

Abbreviations: ED = emergency department; TBT = time-based-targets; IT = information technology; NEAT = National Emergency Access Targets

Six studies were from Australia^51-56^, three from New Zealand^57-59^, three from England^60-62^, and one from Canada^63^. The studies were qualitative^52 53 56 59-63^ or mixed methods^51 54 55 57 64^. The included study settings were all single-payer, government-run health care systems, in English speaking, high-income countries. All studies purposefully recruited participants, representing a wide range of stakeholders. In total 617 interviews were reported. Three studies derived from the same participant interviews in New Zealand^57-59^; four derived from the same participant interviews in Australia^51-54^.

Most participants were emergency and hospital staff, including managers, senior and junior medical and nursing staff, allied health professionals, clerks, and project team members. One study^55^interviewed representatives from medical bodies, and members of the public.

The included studies were generally assessed as being of high quality with low risk of bias. A few studies had a higher risk of bias due to small study populations. The appraisal and risk of bias for each included study is described in Table 3.

**Table 3.** Critical appraisal and risk of bias of included studies.

| Paper | Aims | Methods and design | Recruitment | Data collection | Analysis | Risk of bias |
| --- | --- | --- | --- | --- | --- | --- |
| Forero, 2019<sup>51</sup> | What was the association between 4-hour performance improvements (high, moderate and low) and impact on staff, models of care, communication and clinical outcomes? | Mixed-methods approach to explain quantitative findings of level of performance in meeting target. Methodology and design described in separate paper<sup>65</sup>. | Stratified purposive sampling (quota sampling), criterion-based and maximum variation sampling strategies of hospitals and participants. Eligible participants needed to be working in the ED during the period that the target policy was implemented. No patient voice. | Semi-structured interview technique. Integrated interview protocol and form of data well described. Saturation of data achieved. Ethics approval was obtained and confidentiality of participants assured. | Phenomenological and grounded theory approach to analysis, with triangulation of data, rigorous methods regarding recruitment, interviewing, data checking, analysis and theme saturation; no patient voice. | Low risk of bias. Overall very good methods due to multi-centre, purposive, stratified, representative of range of stakeholders (although not consumers) |
| Forero, 2019<sup>52</sup> | What was the psychosocial impact of the 4-hour policy? What were the intended and unintended consequences of the 4-hour policy described by the Diffusion of Innovation Theory? | Qualitative, multi-centre. Methods described in separate paper<sup>65</sup>. | Stratified purposive sampling (quota sampling), criterion-based and maximum variation sampling strategies of hospitals and participants. Eligible participants needed to be working in the ED during the period that the target policy was implemented. No patient voice. | Semi-structured interview technique. Integrated interview protocol and form of data well described. Saturation of data achieved. Ethics approval was obtained and confidentiality of participants assured. | Phenomenological and grounded theory approach to analysis, with triangulation of data, rigorous methods regarding recruitment, interviewing, data checking, analysis and theme saturation. | Low risk of bias. Overall very good methods due to multi-centre, purposive, stratified, representative of range of stakeholders (although not consumers) |
| Nahidi, 2019<sup>53</sup> | Based on staff experiences, what changes in ED management resulted from the 4-hour rule policy? | Qualitative, multi-centre. Methods described in separate paper<sup>65</sup>. | Stratified purposive sampling (quota sampling), criterion-based and maximum variation sampling strategies of hospitals and participants. Eligible participants needed to be working in the ED during the period that the target policy was implemented. No patient voice. | Semi-structured interview technique. Integrated interview protocol and form of data well described. Saturation of data achieved. Ethics approval was obtained and confidentiality of participants assured. | Phenomenological and grounded theory approach to analysis, with triangulation of data, rigorous methods regarding recruitment, interviewing, data checking, analysis and theme saturation. | Low risk of bias. Overall very good methods due to multi-centre, purposive, stratified, representative of range of stakeholders (although not consumers) |
| Nahidi, 2019<sup>54</sup> | What were the impacts and outcomes perceived by ED staff during the implementation of the 4-hour rule policy in their local ED environment? | Mixed-methods approach to explain quantitative findings of level of performance in meeting target. Methodology and design appropriate and justified. Methods described in separate paper<sup>65</sup>. | Stratified purposive sampling (quota sampling), criterion-based and maximum variation sampling strategies of hospitals and participants. Eligible participants needed to be working in the ED during the period that the target policy was implemented. No patient voice. | Semi-structured interview technique. Integrated interview protocol and form of data well described. Saturation of data achieved. Ethics approval was obtained and confidentiality of participants assured. | Phenomenological and grounded theory approach to analysis, with triangulation of data, rigorous methods regarding recruitment, interviewing, data checking, analysis and theme saturation; no patient voice. | Low risk of bias. Overall very good methods due to multi-centre, purposive, stratified, representative of range of stakeholders (although not consumers) |
| Stokes, 2011<sup>55</sup> | Review of the Four Hour Rule Program (FHRP) in WA. Clear | 4-hour rule program progress and issues review. | Purposive, stratified recruitment of sites and participants, with snowballing at each | Interviews. No description of interview technique or | The report alludes to using qualitative methodology, | High risk of bias. The selection process was arbitrary and highly prone to |
|  | statement of aims, expressed as Terms of Reference. |  | site and self-selected volunteers. The selection process was arbitrary and highly prone to selection bias (people with vested interests or agenda more likely to be included). Stakeholders external to hospitals have been included, including patients and consumer groups. | data saturation. No mention of ethics approval or confidentiality of participants. | with some triangulation of data against alternate sources. There is limited methods description available. | selection bias. |
| Leggat, 2016<sup>56</sup> | What were the perceptions of staff involved in ED process redesign, of the impact of the redesign on the quality of patient care? | Single centre, qualitative study | Participants approached after hospital identified them as being involved in process redesign project using non-probability sampling. Unclear sampling process, regarding both the site and the participants. No patient voice. | Interviews with participants. Ethics approval obtained. | Thematic analysis approach until theme saturation achieved. Three coders, data cross-checking undertaken. No description of relationship between interviewers and participants. | High risk of bias due to small number of respondents in single-centre and recruitment concerns. |
| Tenbenschel, 2020<sup>57</sup> | Regarding ED time-based-target implementation: how and why does gaming vary over time and between organisations? | Mixed-methods approach using comparative case studies to explain quantitative findings of "gaming". Methodology | Purposeful stratified recruitment of participants and hospitals. Number of participants chosen for pragmatic reasons and to achieve saturation. Discussion of selection of | Data collected using semi-structured interviews. Topic guide and form of data well described. Saturation of data achieved. In | In-depth description of analysis approach including how themes/categories were derived from the data. Inductive and deductive | Low risk of bias. Overall very good methods due to multi-centre, purposive, stratified, representative of range of stakeholders (although not |
|  |  | and design appropriate and justified. Methods described in separate paper<sup>66</sup>. | participants and why some people chose not to take part. Note poor response from nursing staff. | a small number of interviews, some participants were not responsive to probes for clarity in relation to sensitive issues. Ethics approval was obtained and confidentiality of participants assured. | approach to analysis, some triangulation of data against surveys. Maori representation on research team. | consumers) |
| Tenbenschel, 2017<sup>58</sup> | Which actions taken by hospitals can account for overall ED LOS reductions? | Mixed-methods approach built on comparative case studies to explain quantitative variation in ED LOS across different sites. Methodology and design appropriate and justified. Methods described in separate paper<sup>66</sup>. | Purposeful stratified recruitment of participants and hospitals. Number of participants chosen for pragmatic reasons and to achieve saturation. Discussion of selection of participants and why some people chose not to take part. Note poor response from nursing staff. | Data collected using semi-structured interviews. Topic guide and form of data well described. Saturation of data achieved. In a small number of interviews, some participants were not responsive to probes for clarity in relation to sensitive issues. Ethics approval was obtained and confidentiality of participants assured. | In-depth description of analysis approach including how themes/categories were derived from the data. Inductive and deductive approach to analysis, some triangulation of data against surveys. Maori representation on research team. | Low risk of bias. Overall very good methods due to multi-centre, purposive, stratified, representative of range of stakeholders (although not consumers). |
| Tenbensen, 2016<sup>59</sup> | An examination and comparison of the implementation consequences of two NZ health targets: immunisation rates for 2 year olds and ED 6-hour-rule policy. | Qualitative research techniques including key-informant interviews and document analysis, within implementation science framework. Methods described in separate paper<sup>66</sup>. | Purposeful stratified recruitment of participants and hospitals. Number of participants chosen for pragmatic reasons and to achieve saturation. Discussion of selection of participants and why some people chose not to take part. Note poor response from nursing staff. | Data collected using semi-structured interviews. Topic guide and form of data well described. Saturation of data achieved. Ethics approval was obtained and confidentiality of participants assured. | In-depth description of analysis approach including how themes/categories were derived from the data. Inductive and deductive approach to analysis, some triangulation of data against surveys. Maori representation on research team. | Low risk of bias. Overall very good methods due to multi-centre, purposive, stratified, representative of range of stakeholders (although not consumers). |
| Weber, 2011<sup>60</sup> | What lessons can be learned by organisations from the 4-hour target implementation to successfully meet the target and avoid unintended negative consequences? | Qualitative, multi-centre. some risk of bias due to high rate of non-participation (15 out of 35 trusts declined to participate) and mainly only used single data analyst. | Purposive, stratified sampling of sites until 9 consented to participate (performance against target and type of hospital). 15 of 35 Trusts declined consent to participate in the larger, stratified, quantitative study, requiring routine ED data. Of those remaining, 11 Trusts were approached until 9 Trusts agreed to participate in qualitative interviews. | Semi-structured interview with open-ended questions. Respondents were assured on anonymity for themselves and their organisation. 25 interviews were face-to-face, 2 were by telephone. | Content analysis was undertaken, to determine themes and examples of themes for each ED. Themes were grouped and their occurrence across sites was compared. Minority perspectives were incorporated. Disagreements between coders were discussed until agreement was obtained. | Moderate risk of bias due to high rate of non-participation (15 out of 35 trusts declined to participate) and mainly only used single data analyst, however otherwise good methodology. |
|  |  |  | No patient voice. |  | Theme saturation was achieved. Rigorous methods were used to synthesize results. |  |
| Mortimore, 2007<sup>61</sup> | What are emergency nursing views on the advantages and disadvantages of the 4-hour target? | Qualitative, single-centre. | Purposive recruitment from a stratified sample of staff, unclear how participants were recruited. No patient voice. | Semi-structured interviews. Ethics approval obtained. Independent interviewer to maintain participant anonymity. No mention of theme saturation. | Inductive approach with framework analysis | High risk of bias due to small number of respondents, recruitment concerns and no theme saturation |
| Vezyridis, 2014<sup>62</sup> | An examination of changes in clinical and organisational processes to meet the 4-hour target, focusing on the role of space, time and information technology to optimise patient flow | Qualitative, single-centre. | Purposive recruitment, inclusion/exclusion criteria unclear, poor methodology description regarding sampling/recruitment, relationships between interviewers and research team. No doctor or patient voice. | Semi-structured interviews. No theme saturation. Ethics approval obtained. | Thematic analysis. Discussion of findings with ED nurse team leader. | High risk of bias due to small number of respondents, recruitment concerns and no theme saturation |
| Rotteau, 2015<sup>63</sup> | A description of hospital-based team experiences | Qualitative, multi-centre | Purposive, stratified recruitment of sites (best and worst amount of | Semi-structured interviews. Thematic saturation | Thematic analysis approach as per Braun and Clarke methods; no | Moderate risk of bias. Multisite project with good sampling of sites; |
|  | during process improvement implementation; team perceptions of factors influencing success or failure of the changes |  | performance change); no description of how individuals were recruited from each site. No patient voice, no non-managerial health care worker voice. | achieved. Ethics approval obtained. | mention of recruitment methodology. It is unclear why only 24 interviews are presented (possibly as theme saturation was achieved). | limited information about individual recruitment methods and a lack of clarity regarding the reason for a smaller sample than predicted to be required. |

We identified 53 outcomes from the included studies and categorised each into a theme. Themes explored perceptions of the impact of time-based-targets on quality and safety of care, timeliness of investigations or treatment, length-of-stay, receiving care, representations, unintended consequences, patient and staff experiences, and enablers and barriers to implementing time-based-targets. For each theme, we have described key findings and our confidence in each finding. A narrative review of each theme is presented below. A detailed analysis is shown in Table 4.

**Table 4.** Assessment of confidence in findings using CERQual

| Summary of review finding | Studies contributing to review finding | Methodological limitations | Coherence | Adequacy | Relevance (Internal validity - how relevant were the included studies to this review) | Overall assessment of confidence in finding |
| --- | --- | --- | --- | --- | --- | --- |
| Quality and safety of ED care:<br>Implementation of TBT could have a positive or negative impact on quality of patient care. Positive experiences were related to improved patient flow and access to timely care, whereas negative experiences occurred when meeting the target was prioritised over patient care. | Forero, 2019<sup>51</sup>;<br>Forero, 2019<sup>52</sup>;<br>Nahidi, 2019<sup>53</sup>;<br>Nahidi, 2019<sup>54</sup>;<br>Stokes, 2011<sup>55</sup>;<br>Leggat, 2016<sup>56</sup>;<br>Tenbensen, 2017<sup>58</sup>;<br>Tenbensen, 2016<sup>59</sup>;<br>Mortimore, 2007<sup>61</sup>;<br>Weber, 2011<sup>60</sup>;<br>Vezyridis, 2014<sup>62</sup>;<br>Rotteau, 2015<sup>63</sup> | Minor concerns. There were 12 studies contributing to this finding, of varying methodological quality ranging from minor to significant concerns. Two studies were judged to be at risk of bias due to concerns about recruitment including selection bias and small number of respondents. However, since there was no discrepancy in outcomes between studies with high and low risk of bias, we have only minor concerns about the methodological limitations of this finding. | No concerns. The data in the primary studies was varied, with respondents reporting positive, negative and neutral experiences with the targets. The finding describes the complexity of data and explores elements that contributed to those perceptions. | Minor concerns. High number of respondents from Australia, NZ, UK and Canada, with a range of different job roles and experiences (although notably not the consumer). | No concerns. The data from the primary studies is directly relevant to the context of the review question. | High confidence due to finding being drawn from large number of respondents and showing high levels of coherence. Some studies have significant methodological concerns yet the finding is consistently supported by studies with only minor methodological concerns. |
| Access block and overcrowding: Clinicians and managers observed improved patient flow and shorter ED stays. However, most EDs did not consistently meet their target, particularly for mental health patients, and over time due to increasing attendances and complexity, independent of the time-based target. There was also concern about unintended consequences of the target such as increasing admissions and representations due to premature discharges, which could potentially increase ED overcrowding. | Forero, 2019<sup>51</sup>; Forero, 2019<sup>52</sup>; Nahidi, 2019<sup>53</sup>; Nahidi, 2019<sup>54</sup>; Tenbensen, 2017<sup>58</sup>; Stokes, 2011<sup>55</sup>; Leggat, 2016<sup>56</sup>; Mortimore, 2007<sup>61</sup>; Weber, 2011<sup>60</sup> | Moderate concerns. Theme best suited to qualitative methodology rather than quantitative. Poor correlation between staff perception on access block and actual level of performance with regards to target. Selection bias may have under-represented "underperforming" health services due to fear of repercussions. | No concerns. The data in the primary studies was varied, with respondents reporting positive, negative and neutral experiences with the targets. The finding describes the complexity of data and explores elements that contributed to those perceptions. | Minor concerns. High number of respondents from Australia, NZ and UK with range of different job roles and experiences, although notably not the consumer. | No concerns. The data from the primary studies is directly relevant to the context of the review question. | Low confidence predominantly due to concern about the appropriateness of qualitative methodology to explain this finding. |
| Patient experience: Clinicians and managers perceived that patients were positive about TBT because of improved access to tests and | Forero, 2019<sup>52</sup>; Stokes, 2011<sup>55</sup>; Leggat, 2016<sup>56</sup>; Mortimore, | Serious concerns. None of the studies were specifically designed to address the question of patient experience. Four of the included | Minor concerns. Although there is limited data on the patient experience, the variation in the | Serious concerns about adequacy of data, due to lack of patient perspective contributing to | No concerns. The data from the primary studies is directly relevant to | Very low confidence, predominantly due to inadequate data representing the patient voice, and methodological limitations of the |
| inpatient beds. However, the trade-off was limited time for ED staff to build rapport and communicate with patients. | 2007<sup>61</sup>; Tenbensen, 2016<sup>59</sup>; Rotteau, 2015<sup>63</sup> | studies had serious methodological limitations. | data is described in the theme. | finding. | the context of the review question. | included studies that increase the risk of bias. |
| Staff morale and workload: Clinicians from emergency and other hospital departments described how pursuit of targets affected their workload and morale at work. In many cases, clinicians experienced increased stress and lowered morale due to increased pressures associated with meeting the target and feelings of being micro-managed. However improved flow and reduced overcrowding could alleviate stress. | Forero, 2019<sup>51</sup>; Forero, 2019<sup>52</sup>; Nahidi, 2019<sup>53</sup>; Nahidi, 2019<sup>54</sup>; Stokes, 2011<sup>55</sup>; Vezyridis, 2014<sup>62</sup>; Tenbensen, 2016<sup>59</sup>; Tenbensen, 2020<sup>57</sup> | Very minor concerns only. Although some papers had methodological limitations, these limitations were not perceived to bias confidence in this finding, due to no discrepancy in outcomes between studies with high and low risk of bias. | No concerns. The data in the primary studies was varied, with respondents reporting positive, negative and neutral experiences with the targets. The finding describes the complexity of data and explores elements that contributed to those perceptions. | No concerns. High number of respondents from Australia, NZ and the UK with range of different job roles and experiences. | No concerns. The data from the primary studies is directly relevant to the context of the review question. | High confidence, due to no or very minor concerns about methodology, coherence, adequacy and relevance. |
| Intrahospital and interdepartmental relationships: Relationships between ED and other hospital departments improved when a whole-of- | Forero, 2019<sup>51</sup>; Forero, 2019<sup>52</sup>; Nahidi, 2019<sup>53</sup>; Nahidi, | Very minor concerns only. Although some papers had methodological limitations, these limitations were not perceived to bias | No concerns. The data in the primary studies was varied, with respondents reporting | No concerns. High number of respondents from Australia, NZ and the UK with range of different job | No concerns. The data from the primary studies is directly relevant to | High confidence, due to no or very minor concerns about methodology, coherence, adequacy and relevance. |
| hospital approach to implementing targets was adopted but deteriorated where there was lack of buy-in from non-ED clinicians and management. | 2019<sup>54</sup>; Stokes, 2011<sup>55</sup>; Leggat, 2016<sup>56</sup>; Tenbensen, 2017<sup>58</sup>; Tenbensen, 2016<sup>59</sup>; Vezyridis, 2014<sup>62</sup> | confidence in this finding, due to no discrepancy in outcomes between studies with high and low risk of bias. | positive, negative and neutral experiences with the targets. The finding describes the complexity of data and explores elements that contributed to those perceptions. | roles and experiences. | the context of the review question. |  |
| Clinical education and training: Pursuit of the TBT reduced opportunities for clinical education and training for ED nurses and junior doctors, except where implementation involved closer working relationships between junior and senior doctors. | Forero, 2019<sup>51</sup>; Nahidi, 2019<sup>53</sup>; Nahidi, 2019<sup>54</sup>; Stokes, 2011<sup>55</sup>; Mortimore, 2007<sup>61</sup>; Weber, 2011<sup>60</sup> | No or minor concerns. Although some papers had methodological limitations, these limitations were not perceived to bias confidence in this finding, due to no discrepancy in outcomes between studies with high and low risk of bias. | No concerns. The data in the primary studies was varied, with respondents reporting positive, negative and neutral experiences with the targets. The finding describes the complexity of data and explores elements that contributed to those perceptions. | Serious concerns about adequacy of data, due to lack of junior doctor perspective contributing to finding. | No concerns. The data from the primary studies is directly relevant to the context of the review question. | Very low confidence due to inadequate data representing junior doctor perspectives. |
| Use of time-based targets induces gaming: Clinicians and managers observed distorted responses in order to meet the target, such as diverting patients to short-stay units when ward beds weren't available. | Nahidi, 2019<sup>53</sup>; Nahidi, 2019<sup>54</sup>; Tenbense, 2017<sup>58</sup>; Tenbense, 2016<sup>59</sup>; Tenbense, 2020<sup>57</sup> | No concerns due to high quality methodology in the included studies. | Minor concerns due to Australian studies not indicating to what extent or frequency gaming occurs. NZ studies indicate that gaming is common and widespread. | Minor concerns about quantity and richness of data in Australian studies regarding this finding, and lack of data from English and Canadian perspectives. | No concerns. The data from the primary studies is directly relevant to the context of the review question. | Moderate confidence. Downgraded due to concerns about coherence, adequacy and relevance of data, given that the data only represented narrow contexts. |
| Enablers and barriers to successful achievement of TBT: Sites that successfully implemented TBT required significant financial, staffing and logistical resources, including a whole-of-hospital approach, top-down buy-in from hospital management and careful attention to change management. Where these were not provided, TBT were perceived negatively. | Forero, 2019<sup>51</sup>; Forero, 2019<sup>52</sup>; Nahidi, 2019<sup>53</sup>; Nahidi, 2019<sup>54</sup>; Stokes, 2011<sup>55</sup>; Leggat, 2016<sup>56</sup>; Tenbense, 2020<sup>57</sup>; Tenbense, 2017<sup>58</sup>; Tenbense, 2016<sup>59</sup>; Mortimore, 2007<sup>61</sup>; Weber, 2011<sup>60</sup>, | Very Minor concerns only. Although some papers had methodological limitations, these limitations were not perceived to bias confidence in this finding, due to no discrepancy in outcomes between studies with high and low risk of bias. | No concerns. The data in the primary studies was varied, with respondents reporting positive, negative and neutral experiences with the targets. The finding describes the complexity of data and explores elements that contributed to those perceptions. | No concerns. High number of respondents from Australia, NZ, UK and Canada with range of different job roles and experiences. | No concerns. The data from the primary studies is directly relevant to the context of the review question. | High confidence, due to no or very minor concerns about methodology, coherence, adequacy and relevance. All studies contributed data on this finding and demonstrated both consistency and nuance in findings. |
|  | Vezyridis,<br>2014<sup>62</sup>;<br>Rotteau,<br>2015<sup>63</sup> |  |  |  |  |  |

### quality and safety of ED care

Overall 12 studies contributed to this theme^51-56 58-63^. We have high confidence in this finding due to a large number of participants, representing a wide range of settings and perspectives. Implementation of time-based-targets for emergency length-of-stay had both positive and negative impacts on the capacity to deliver emergency care.

Respondents reported positive experiences when targets were used as a lever to improve patient flow and timely care. Clinicians and managers who perceived that the introduction of targets had a positive impact on quality and safety of care noted that pursuit of the time-based-targets led to improved processes. These allowed emergency nurses and physicians to work more efficiently, refer earlier and perform fewer tests, leading to expedited delivery of patient care ^55 61 62^. In these facilities, sources of delay were investigated and hospitals invested in quality improvement activities to meet the targets, such as new infrastructure and new staffing roles^54 62 63^. One Australian emergency physician observed:

> *“It probably has a positive* [*impact*] *in that we’re making decisions and getting patients to the ward earlier…”*^*51*^

In England, an emergency clinician noted:

> *“Before the target, people were stacked in the corridors on trolleys. Care is better because we have got rid of the corridors of shame*.*”*^*60*^

Negative experiences occurred when targets were prioritised over patient needs. Health staff in all roles reported this issue^53 55 56 62^. Hasty decision-making impacted negatively on patient assessments, increased test ordering and increased misguided care by junior doctors^53^. Allied health staff reported that medical patient evaluations were less detailed^59^. Clerical staff administration errors also increased; which was attributed to productivity pressures^59^.

> *“…somebody’s in pain and you need to get them on the ward within those 4 hours - do you move them or not?” Emergency nurse, England*^*61*^

There was a perception of increased patient churn and a chaotic environment in wards, which made it more difficult for clinicians to maintain quality of care, exposing patients to increased risk^54^. Ward staff felt they were over-census at times, reducing patient privacy and safety^59^.

### access block and overcrowding

Nine studies contributed to this theme^51-56 58 60 61^. We have low confidence in this finding as quantitative methodology is better suited to exploring this theme, leading to moderate methodological concerns about this finding.

Many emergency staff and managers found that patient flow and access block improved with the targets^51 53 55 61^even if they did not meet the target. This was attributed to process changes driven by the creation of a closely monitored target^56 59^. Different hospitals used different strategies to meet targets. Some sites implemented rapid assessment models, streaming to short-stay units, use of new coordinator roles and improved discharge processes. These changes had varying success in improving access block and ED overcrowding. There is insufficient data from these studies to comment on which models of care are most successful in reducing access block and overcrowding. Targets didn’t improve access block for psychiatric patients in Australia^55^. New models of care were found in hospitals in all settings and geographical locations. They were implemented at a local level and there was significant variation in the approach to meeting targets.

Emergency staff were concerned that the policy could worsen emergency overcrowding through increased emergency attendances (because consumers thought there would be shorter queues as a result of the policy)^61^, increased representations (having discharged patients too early)^59^ and higher admission rates leading to reduced inpatient bed capacity^55 56^. These concerns were disproven in a quantitative systematic review on the topic (*Under review, BMJ Open*).

### patient experience

Six studies contributed to this finding^52 55 56 59 61 63^. We have very low confidence in this finding due to inadequate data representing the patient voice. Only one study includes a consumer perspective.

In Western Australia, feedback from the Health Consumers’ Council was very positive^55^. Clinicians in New Zealand, England, and Canada reported fewer patient complaints^59 63^ and felt that patient satisfaction increased^61^. Some clinicians were concerned that the targets reduced the capability of emergency staff to build rapport with patients^52^.

### staff morale and workload

Eight studies contributed to this finding^51-55 57 59 62^. We have high confidence in this finding because the contributing data represents a wide range of settings and perspectives. While some of the included studies had methodological limitations, there was agreement between findings from these studies compared to others that were assessed as being of high methodological quality.

Clinicians reported a range of positive and negative experiences associated with the introduction of targets. Many doctors and nurses experienced increased stress and lowered morale due to increased pressures associated with meeting the target. For example, one Australian emergency director felt that:

> *“all you’re saying to the staff is, ‘You’re not good enough. You don’t work hard enough. You can’t do your job fast enough’”*^*51*^

They also described a sense that they were being micro-managed and under surveillance^51 53 59 61 62^. More than one respondent describing feeling like they were being watched by ‘Big Brother’^54 61^. Many participants reported an increase in their workload in relation to the targets, including increased documentation requirements^52 53^ and reduced efficiency due to ‘workarounds’^62^. This had the effect of reducing nursing morale and increasing staff turnover, with one nurse reporting that junior nurses were leaving:

> *“because they feel they cannot cope with high pressure work all the time”*^61^

Furthermore, patient awareness of the target could increase stress for emergency nurses because patients felt enabled to challenge delays^61^. Junior doctors reported feeling bullied by nurses allocated to the role of time-based-target compliance^51 52 55 62^. Ward staff felt increased pressure to discharge patients from hospital wards^59^.

Conversely, some emergency staff reported less stress, more control over their environment and an enhanced ability to manage increased attendances^52 53^. This was attributed to improvements in staffing and resources, increased control over the admission process and improved autonomy for emergency medicine regarding decision-making^60 62^. Administration staff morale and stress didn’t change in Australia^51-54^.

### intrahospital and interdepartmental relationships

Nine studies contributed to this finding^51-56 58 59 62^. We have high confidence in this finding because the contributing data represents a wide range of settings and perspectives. While some of the included studies had methodological limitations, both higher and lower quality studies presented similar findings.

Time-based-targets impacted on relationships and the interface between emergency medicine, other hospital departments and hospital leadership. At many sites, the emergency department was deemed responsible and accountable for delivering targets, despite a lack of hospital system and process change within the hospital, which exacerbated intra-hospital tensions^52 59 60^.

#### For example

> *“Oh that’s your 4-hour target” UK emergency nurse trying to move a patient to the ward who was close to a 4-hour emergency stay*^*60*^

Similarly, another emergency nurse in Australia stated:

> *“We had to harass the wards staff which had a negative throwback towards us, and we were harassed by executives, nursing and others, who would come down on a regular basis to explain and justify why patients had not left the emergency department”* ^51^

There was resistance to change and poor engagement from inpatient junior doctors and specialists who perceived that the targets didn’t involve themS^55 58 59^.

Conversely, some emergency staff found improved relationships with inpatient units, as responsibility for the care of acutely unwell patients shifted to the whole hospital^53 59^. Having a target could enhance professional relationships and interactions with the rest of the hospital when it provided a common purpose and was managed collaboratively and collectively^52 58 67^.

There was no association between jurisdiction, specific target, or staff role as to whether targets were perceived to have a positive or negative impact on intra-hospital relationships. The most plausible explanation for this variance is that relationships improved in hospitals where there was a “whole of hospital approach” and deteriorated when the target was applied in a punitive way that placed responsibility on the ED to meet the target, without considering the interface with other departments and services.

### clinical education and training

Six studies contributed to this finding^51 53-55 60 61^. We have very low confidence in this finding due to inadequate data from junior doctors.

Emergency clinician and managers felt that medical education training environments had deteriorated, with fewer training opportunities and less time for junior doctors and nurses to learn^53-55 60 61^;. Hospital managers deprioritised training and research, compared to targets^53^. One Australian emergency physician observed:

> *“I think the biggest impact that it’s had is that it’s actually sunk all our resources into trying to achieve that one target so that other aspects of our normal work which we think are important, such as education and research, are just continually being eroded”*^*51*^

Some participants reported that role restructuring in order to meet the targets led to greater emergency team integration, having a positive impact on medical education, and improved educational opportunities^53^. Another Australian emergency physician reported:

> *“There is very strong evidence that actually working with the junior doctors is actually better teaching than letting them bumble through it”*^55^

In the UK, a senior clinician described both the positive and negative impacts of targets on training and education:

> *“In general, the target has had a deleterious effect* [*on training*] *not as much opportunity to stand and talk… It takes them twice as long to do procedures—so we are not always able to let them do these. However, what was training like when we had 80 patients in the department?”*^60^

### gaming

Five studies contributed to this finding^53 54 57-59^. We have moderate confidence in this finding as the data comes from New Zealand and Australian settings only.

Gaming has been described as *“making performance appear better when it is not”*^57 66^ Clinicians and managers found that gaming occurred in order to meet targets^53 54 58 59^. This included avoiding target breaches by diverting patients to short-stay units when ward beds weren’t available^58^. This behaviour is described by one Western Australian emergency nurse as:

> *“The patient is about to breach, just get them into a bed, any bed, instead of taking a little bit longer to make an accurate disposition decision”*^*55*^.

Gaming was universal and occurred more frequently when there was pressure from senior hospital management staff on ED or the “whole-of-hospital” to meet the target, but at lower levels when ED staff did not feel pressure to meet the target^57^.

### enablers and barriers to achieving targets

Thirteen studies contributed to this finding51-63. We have high confidence in this finding because the data reflected a wide range of settings and perspectives, with a high level of consistency in outcomes.

There was a high degree of consensus that successfully achieving the targets required a top-down and whole-of-hospital approach^51 58 60 63 67^. Describing the targets as ‘emergency time-based-targets’ was felt to be misguided, as efforts to improve timeliness were impacted by processes that involve the whole hospital system^55 56 62^.

Factors that were thought to be required for successful change management included careful pre-planning, staff education and changes tailored to the local environment^54^, provision of short-term project support officers55 63, learning from other sites59 and executive and managerial buy-in^58^.

Hospitals introduced a broad range of new processes and models of care. These included changes to the handover process between emergency and inpatient units^54^ and new targets for patient work-up and ward bed allocation58. Improvements in out-of-hours discharge planning and patient discharge lounges were helpful58. Extended opening hours support services (e.g. diagnostics and pharmacy) was also required61

Barriers to target achievement included lack of ED authority regarding admissions and insufficient support from logistics or other areas of the hospital^54^. Pursuit of diversion strategies was largely ineffective^58^. A need to engage community referrers in future acute care service planning was reported61. Financial resources and physical changes in capacity and layout were required to implement the changes. Without this, sites had inadequate resources to implement the policy or manage the workload^53-55 62^.

Hospitals in New Zealand rostered more doctors, nurses and orderlies in both emergency departments and wards^58^. In England, emergency nurse practitioner numbers increased to see “minor” patients more efficiently262. The skill mix of emergency clinicians and inpatient ward staff was identified as important, with a need for more senior and experienced clinicians to be involved earlier in the course of the patient journey^54 55 60^. Hospitals upgraded communications and information technology systems to meet time-based-targets, at times improving flow management, but at other times reducing efficiency^54 58 62^.

### overall

The variation in experience was not dependent on jurisdiction, country or time target or threshold with respondents from hospitals across Australia, NZ, Canada and the UK all contributing both positive and negative impressions, experiences and impacts on care quality. These experiences were universal across jurisdictions. The impact of targets was associated with how the various targets were implemented and achieved. When applying targets, positive effects on quality of care were noticed when health services adopted a whole-of-hospital response and ensured that meeting the target did not occur at the expense of patient care activities, or training for junior clinicians. Negative effects were experienced by staff when the target was prioritised over patient care, there wasn’t whole hospital engagement and when resources weren’t adequate to implement and maintain target achievements (such as physical layout and capacity, increased staffing, processes, change management personnel, information technology support, inpatient beds, and inpatient engagement).

## DISCUSSION

This qualitative systematic review assessed the impact of time-based-targets for emergency patient length-of-stay on quality of care. The review included thirteen studies from four different countries and settings, including 617 participant interviews from multiple hospitals. The eight themes were: quality and safety of care, access block and overcrowding, patient experience, staff workload and morale, intra-hospital and interdepartmental relationships, clinical education and training, gaming behaviours, and barriers and enablers to implementing targets.

This is the first review to compare and contrast experiences of targets across different settings. In this review, clinicians described common experiences of targets across different countries and target times. For example, clinicians from Australia, NZ, and Canada, operating under target times of 4, 6 or 8 hours, identified situations where meeting the target took priority over patient care. However, within the same jurisdiction and targets, clinicians also had very different experiences of the target. For example, Australian clinicians operating under a 4-hour target reported variously that their workload, stress, and relationships with inpatient colleagues improved or deteriorated. This suggests that the impact of targets does not relate to the policy itself or even the time frame, but rather local factors that influenced how the policy was implemented and perceived. This interpretation is supported by the findings of the quantitative systematic review, which demonstrated considerable heterogeneity in findings between studies at different sites (*Under review, BMJ Open*).

The strengths of this study are its comprehensive approach and robust methodology. This helps to mitigate concerns that qualitative studies lack generalisability to other settings. The findings of this review are likely to be generalisable to health systems that are similar to those in Australia, NZ, the UK and Canada. None of the studies were from fee-for-service environments, nor from low or middle-income countries or non-English speaking regions and caution should be used in applying these findings to other settings.

Weaknesses of this study include limited input from consumers into the studies. Very few studies report the impact of time-based-targets on inpatient wards or primary care. Three studies were small, single-site evaluations with limited methodology. We have very low confidence in the findings regarding patient experience or clinical education and training. We have low confidence in findings regarding access block and overcrowding.

Time-based-targets are a policy tool that aims to improve quality of care. Targets can improve patient flow, efficiency and interdepartmental relationships. In order to be effective however, targets should be implemented using a collaborative approach that is driven from the top down, ensuring that no one department is under pressure to meet the target without adequate resourcing and support. Activities driven by the target should be adequately funded to ensure that staff are not forced to compromise on patient care or non-clinical activities (such as education and training) in order to meet the target.

Further research should address which target and incentives to adopt; economic implications of targets; the impact on hospital resources and elective care; and the impact of targets in different settings. Research questions should include patient perspectives and patient-centred questions about care delivery and processes.

## conclusion

Time-based-targets for emergency length-of-stay have impacted on the quality of emergency patient care. The impact is varied, can be both positive and negative and successful implementation depends on whole hospital resourcing and engagement with targets.

## Data Availability

All data is publicly available already.

## contributor and guarantor information

Study design: PJ, KW, BH; Design revision: all authors; Literature Search: BH; Abstract screening: DH, KW, DM, VG, EM, PM, RM, PJ, YN; Data Extraction: KW, BH, PJ, VG, DH, DM, RM; GRADE: KW, BH; Write-up: KW, BH, PJ; Revisions: All authors.

## conflicts of interest

All authors have completed the ICMJE uniform disclosure form at www.icmje.org/coi_disclosure.pdf and declare: organisational support for the review from the Australasian College for Emergency Medicine. No financial support was received for the submitted work. Many of the authors work in emergency departments in Australia and New Zealand, which have been subject to time-based-targets. Peter Jones, Roberto Forero and David Mountain published original research manuscripts on the topic, some of which have been included in the review. They have also received government and emergency college funding for time-based-target research.

## Notes

### Clinical Protocols

https://www.crd.york.ac.uk/prospero/display_record.php?RecordID=107755

### Funding Statement

No external funding was received.

### Author Declarations

IRB approval not required, no patient information was required for this study.

