## Appendix 1 for "Has the implementation of time-based-targets for emergency department length-of-stay influenced the quality of care for patients? A systematic review of qualitative literature"

**Appendix 1. Search Strategies**

Ovid MEDLINE database search to 19 December, 2018

| 1 | emergency service, hospital/ or emergencies/ or emergency medical services/ or emergency services, psychiatric/ or emergency medicine/ or emergency nursing/ | 147691 |
| --- | --- | --- |
| 2 | ("accident and emergency" or "accident & emergency" or emergency service? or (emergency adj2 (care or healthcare or department? or unit or units or room? or treatment?))).ab,ti. | 116191 |
| 3 | 1 or 2 | 209573 |
| 4 | (quality assurance, health care or "quality of health care" or "health care quality, access, and evaluation" or quality indicators, health care or quality improvement or "Outcome Assessment (Health Care)" or evaluation studies or program evaluation).sh. | 484592 |
| 5 | (target? or rule? or standard? or strateg* or performance or require* or complian* or breach* or delay* or throughput).ab,ti. | 5081053 |
| 6 | (access* or wait* or "time based" or "LOS" or "NEAT" or (6 hr or 6 hour or six hour) or (4 hr or 4 hour or four hour)).ab,ti. | 556071 |
| 7 | (length of stay or time or time factors or health services accessibility or patient care management or national health programs).sh. | 1310978 |
| 8 | ("National Emergency Access Target" or "West Australian Emergency Access Target" or "WEAT" or "Emergency Treatment Performance" or "ETP" or shorter stay*).ab,ti. | 1725 |
| 9 | 3 and 4 and 5 and 6 | 856 |
| 10 | 3 and 4 and 5 and 7 | 1340 |
| 11 | 3 and 4 and 8 | 18 |
| 12 | 9 or 10 or 11 | 1769 |

CINAHL database search to 9 January 2019

| S1 | (MH “Emergency Nursing”) | 13748 |
| --- | --- | --- |
| S2 | (MH “Emergency Medicine”) | 10,278 |
| S3 | (MH “Emergency Medical Services”) | 87,209 |
| S4 | TX (“accident and emergency” OR “accident & emergency”) OR TI “emergency department” | 28,205 |
| S5 | S1 or S2 or S3 or S4 | 113,387 |
| S6 | (MH “Quality of Health Care”) | 640,809 |
| S7 | (MH “performance Measurement systems”) OR (MH “Audit”) OR (MH “Nursing Audit”) OR TX (“clinical audit” or “medical audit”) | 20,297 |
| S8 | TX (quality N2 (indicator* or criteri* or standard* or norm*)) | 35,803 |
| S9 | TX (performance N2 (indicator* or measure* or data or rating* or information)) | 38,101 |
| S10 | S6 OR S7 OR S8 OR S9 | 698,635 |
| S11 | TX (Time* or wait* or throughput or “length of stay” or “shorter stay” or “emergency”) N3 (target*)) | 7,587 |
| S12 | TX ((4-hour* or “4 hour” or 4-hr or “4 hr” or 4-h or “4 h” or 4h or four-hour* or “four hour*”) N2 (target* or rule or standard or wait* or delay* or throughput)) | 1,214 |
| S13 | TX ((6-hour* or “6 hour” or 6-hr or “6 hr” or 6-h or “6 h” or 6h or six-hour* or “six hour*”) N2 (target* or rule or standard or wait* or delay* or throughput)) | 340 |
| S14 | TX ((“emergency room” or “emergency care” or “emergency department”) N3 (target* or wait* or “length of stay” or “wait time*”) or throughput or “shorter stay”) | 1,768 |
| S15 | TX ((wait-time or “wait* time”) N3 “strateg*)) | 250 |
| S16 | TX (“emergency access target*” OR “emergency treatment performance” OR “access to emergency care” OR “NEAT” OR “WEAT”) | 3,162 |
| S17 | S11 OR S12 OR S13 OR S14 OR S15 OR S16 | 13,328 |
| S18 | S5 AND S10 AND S17 | 702 |

ABI Inform Proquest database search to 18 Jan 2019

| 1 | AB,TI((target? OR rule? OR standard? OR strateg* OR performance OR require* OR complian* OR breach* OR delay* OR throughput))  AND  AB,TI((access* OR wait* OR "time based" OR "LOS" OR "length of stay" OR "NEAT" OR (6 hr OR 6 hour OR six hour) OR (4 hr OR 4 hour OR four hour)))  AND  AB,TI((emergency AND (department OR room OR service OR unit))  Restricted to 1/1/2000 onwards | 4,889 |
| --- | --- | --- |
| 2 | Apply filter to search by Subjects  Include:  hospitals OR emergency medical care OR emergency services OR health care policy OR health services OR health care access OR health care OR emergency service, hospital OR quality of care OR patient satisfaction OR health care delivery OR efficiency, organizational OR compliance OR emergency medical services OR patient safety OR health services accessibility OR public safety OR health care expenditures OR mortality OR quality OR length of stay OR clinical outcomes OR quality of service  Exclude:  equity stake AND capital formation AND private equity AND securities analysis AND securities offerings AND venture capital AND acquisitions & mergers AND statistical data AND corporate profiles AND petroleum industry AND electric power plants AND wireless networks AND telecommunications industry AND acquisitions \\\\& mergers AND network security AND manufacturing | 818 |

Emerald database search to 18 January 2019

| 1 | Emergency time target  OR  Emergency hour rule  OR  Emergency shorter stay  OR  Emergency wait time | Anywhere in article | 18,913 |
| --- | --- | --- | --- |
| 2 | 1 | Restricted to 1/1/2000 onwards | 14,318 |
| 3 | 2 | Restricted to “Health Services” keyword | 206 |
| 4 | 2 | Restricted to “Health care” keyword | 185 |
| 5 | 2 | Restricted to “Hospitals” keyword | 167 |
| 7 | 2 or 3 or 4 or 5 |  | 460 |

Cochrane database search to 15 January 2019

| #1 | ((target? or rule? or standard? or strateg* or performance or require* or complian* or breach* or delay* or throughput)):ti,ab,kw | 361017 |
| --- | --- | --- |
| #2 | ((access* or wait* or "time based" or "LOS" or "length of stay" or "NEAT" or (6 hr or 6 hour or six hour) or (4 hr or 4 hour or four hour))):ti,ab,kw | 101844 |
| #3 | (("National Emergency Access Target" or "West Australian Emergency Access Target" or "WEAT" or "Emergency Treatment Performance" or "ETP" or shorter stay*)):ti,ab,kw | 5301 |
| #4 | (quality or outcome or improvement or evaluation or assessment):ti,ab,kw | 515306 |
| #5 | ("accident and emergency" or "accident & emergency" or "emergency department*" or "emergency room*" or "emergency unit*" or "emergency medicine" or "emergency nursing" or "emergency service"):ti,ab,kw | 10087 |
| #6 | #1 and #2 | 43951 |
| #7 | #4 and #5 and #6 | 971 |
| #8 | #4 and #5 and #3 | 129 |
| #9 | #7 or #8 | 1030 |

Embase database search to 9 January 2019

| 1 | emergency service, hospital/ or emergencies/ or emergency medical services/ or emergency services, psychiatric/ or emergency medicine/ or emergency nursing/ | 125994 |
| --- | --- | --- |
| 2 | ("accident and emergency" or "accident & emergency" or emergency service? or (emergency adj2 (care or healthcare or department? or unit or units or room? or treatment?))).ab,ti. | 159181 |
| 3 | 1 or 2 | 249464 |
| 4 | (quality assurance, health care or "quality of health care" or "health care quality, access, and evaluation" or quality indicators, health care or quality improvement or "Outcome Assessment (Health Care)" or evaluation studies or program evaluation).sh. | 11787 |
| 5 | (target? or rule? or standard? or strateg* or performance or require* or complian* or breach* or delay* or throughput).ab,ti. | 5724228 |
| 6 | (access* or wait* or "time based" or "LOS" or "NEAT" or (6 hr or 6 hour or six hour) or (4 hr or 4 hour or four hour)).ab,ti. | 645955 |
| 7 | (length of stay or time or time factors or health services accessibility or patient care management or national health programs).sh. | 372962 |
| 8 | ("National Emergency Access Target" or "West Australian Emergency Access Target" or "WEAT" or "Emergency Treatment Performance" or "ETP" or shorter stay*).ab,ti. | 3390 |
| 9 | 3 and 4 and 5 | 166 |
| 10 | 3 and 4 and 6 | 64 |
| 11 | 3 and 4 and 7 | 25 |
| 12 | 3 and 4 and 8 | 0 |
| 13 | 9 or 10 or 11 or 12 | 213 |
