## Appendix 2 for "Has the implementation of time-based-targets for emergency department length-of-stay influenced the quality of care for patients? A systematic review of qualitative literature"

**Appendix 2. Data collection tool template, quality appraisal**

| **Background data** |  |  |  |
| --- | --- | --- | --- |
| Reviewer Name |  |  |  |
| Date |  |  |  |
| Study Name |  |  |  |
| Journal |  |  |  |
| Year of publication |  |  |  |
| **Study details** |  |  |  |
| sponsorship source |  |  |  |
| country |  |  |  |
| setting |  |  |  |
| comments |  |  |  |
| **Author's contact details** |  |  |  |
| Lead author's name |  |  |  |
| Institution |  |  |  |
| email |  |  |  |
| address |  |  |  |
| **Additional identification data** |  |  |  |
| comments |  |  |  |
| **Methods** |  |  |  |
| Design |  |  |  |
| Additional methods data |  |  |  |
| **Population** |  |  |  |
| inclusion criteria |  |  |  |
| exclusion criteria |  |  |  |
| comments |  |  |  |
| **Additional population data** |  |  |  |
| source of data |  |  |  |
| **Baseline characteristics of setting** |  |  |  |
| eg which TBT |  |  |  |
| other comments |  |  |  |
| **Intervention description** |  |  |  |
| Month/Year introduced |  |  |  |
| Details of target |  |  |  |
| incentivized or not |  |  |  |
| details of incentives (carrots/sticks) |  |  |  |
| **Outcomes** | **possibly positive** | **possibly negative** | **neutral** |
| 1 |  |  |  |
| 2 |  |  |  |
| 3 |  |  |  |
| 4 |  |  |  |
| 5 |  |  |  |
| 6 |  |  |  |
| 7 |  |  |  |
| 8 |  |  |  |
| 9 |  |  |  |
| 10 |  |  |  |
| describe as needed eg positive for staff, negative for patients etc. |  |  |  |
| **A. Are the results valid?** |  |  |  |
| 1. Was there a clear statement of aims of the research? |  | Hint: what was the goal of the research? Why was it thought to be important? What was its relevance? |  |
| comments |  |  |  |
| 2. Is the qualitative methodology appropriate? |  | Hint: Does the research seek to interpret or illuminate the actions or subjective experiences of research participants? Is qualitative research the best method for addressing their goal? |  |
| comments |  |  |  |
| **B. Is it worth continuing with the review?** |  |  |  |
| 3. Was the research design appropriate to address the aims of the research? |  | Hint: has the researcher has justified the design? Have they discussed how they came to decide on their methods? |  |
| comments |  |  |  |
| 4. Was the recruitment strategy appropriate to the aims of the research? |  | Hint: has the researcher explained how participants were selected and are these participants appropriate to the question? Have the researchers explained why some participants declined? |  |
| comments |  |  |  |
| 5. Was the data collected in a way that addressed the research issue? |  | Hint: Is the setting justified, is it clear how data was obtained (face-to-face, focus group, structured or semi-structured interviews etc.), are the methods explicit, have the methods been modified mid-study, why? Is the form of data clear? Has theme saturation been discussed and achieved? |  |
| comments |  |  |  |
| 6. Has the relationship between researcher and participants been adequately considered? |  | Hint: has the researcher adequately addressed the potential bias introduced by their role and setting, including recruitment, data collection? Have the researchers considered any change in methodology implications for potential bias? |  |
| comments |  |  |  |
| **C. What are the results?** |  |  |  |
| 7. Have ethical issues been taken into consideration? |  | Hint: are their sufficient details to understand if the research was conducted ethically? How has informed consent been handled? Has ethics committee approval been obtained? |  |
| comments |  |  |  |
| 8. Was the data analysis sufficiently rigorous? |  | Hint: Is there an in-depth description of the analysis process? If thematic analysis was used, was the categoration derivation described? How was data selected from the original sample? Is sufficient data presented to demonstrate results? How are contradictory data presented? Have the authors critically examined their own potential influence on the results? |  |
| comments |  |  |  |
| 9. Is there a clear statement of findings? |  | Hint: are the findings explicit? Is there adequate discussion of the evidence for and against the researchers' findings? Have the researchers discussed the credibility of their findings eg more than one data analyst, triangulation, respondent variation? Are the findings discussed in relation to the original question? |  |
| comments |  |  |  |
| **D. Will the results help locally? Internationally?** |  |  |  |
| 10. How valuable is the research? |  | Hint: does this contribute to a greater understanding of the topic? Do the authors identify where more research is needed? Is there external generalisability/validity? |  |
